# Integrating a conceptual consent permission model from the informed consent ontology for software application execution

**DOI:** 10.1101/2025.01.31.25321503

**Authors:** Muhammad “Tuan” Amith, Yongqun He, Elise Smith, Marceline Harris, Frank Manion, Cui Tao

## Abstract

We developed a simulated process to show a software implementation to facilitate an approach to integrate the Informed Consent Ontology, a reference ontology of informed consent information, to express implicit description and implement conceptual permission from informed consent life cycle. An early study introduced an experimental method to use Semantic Web Rule Language (SWRL) to describe and represent permissions to computational deduce more information from the Informed Consent Ontology (ICO), demonstrated by the use of the All of Us informed consent documents. We show how incomplete information in informed consent documents can be elucidated using a computational model of permissions toward health information technology that integrates ontologies. Future goals entail applying our computational approach for specific sub-domains of the informed consent life cycle, specifically for vaccine informed consent.

## Introduction

In interventional clinical science studies and many health science studies more broadly, there is a process to attain permission from the human participant. In certain cases, this also includes any further studies that utilizes data and information generated by the study for secondary analysis. This process referred to as *informed consent* is defined as the process in which detailed information about a study is presented to the participant in a way that they can understand the general goals, risks and benefits of the study. This understanding helps the participant to agree to the study in a voluntary manner. The informed consent requirements are further elaborated by the Common Rules as part of the U.S. Department of Health and Human Services regulations for the protection of human subjects in research from 45CFR 46^1^. Furthermore, Common Rules is drawn from the Belmont Report ^2^ that provides a framework for ethical guidelines for human research.

Vajda and associates describe the informed consent process as part of the *informed consent life cycle*. The *informed consent life cycle* ``involves not only documentation of the consent when originally obtained, but actions that require clear communication of permission from the initial acquisition of data and specimen through handoffs to, for example, secondary researchers, allowing them access to data or bio-specimens referenced in the terms of the original consent’’^3^. This life cycle connects various permissions from different participating entities, artifacts, data and information, roles and actions, etc. under one unifying umbrella. The menagerie of heterogeneous entities and their changing states introduces management challenges for potential information and communication technology for health care, including relevance in the *learning health system*.

Ontologies aim to model real world entities and provide a standard consensus-agreed representation in a computable, machine-readable format. Potentially, ontologies can represent and manage the heterogeneous entities across the informed consent life cycle. Introduced in 2015, the Informed Consent Ontology (ICO) fulfills this aim with comprehensive coverage of the informed consent process^4^. ICO is manifested as an OWL2 artifact, enabling the possibility to be utilized in software systems. Similar and familiar formal reference ontologies like the Gene Ontology ^5^ and SNOMED CT^6^ have been integrated and impacted clinical software systems (e.g. electronic health care records), and enhanced analytical methodologies for health data^7^. Past studies ^3,4,8^ have highlighted the knowledge engineering and representational focus of ICO, however, very little focus on the application, with the exception of one past study ^9^.

That past study revealed the broad issue of incomplete legal information, including informed consent information imparted to subject participants in research studies. Machine-based reasoning activated from the knowledge base and the predicate logic of an OWL2 ontology could formulate the missing information from informed consent data, which is needed to adhere to ethical requirement of completely informing patients and participants before undergoing participation. That work introduced an attempt to translate data and information from informed consent documents to a computable representation framed by the Informed Consent Ontology and a theoretical permission model (see Section ***Permission Model*** for detailed treatment). Sample permissions from the All of Us documents and Semantic Web Rule Language (SWRL)^10^ rules were encoded onto the Informed Consent Ontology. The generation of the missing information was facilitated by the SWRL rules which produced implicit knowledge from data if the participants agree to engage in the researchers’ study.

### Research Objective

We demonstrate an implementation of how the combination of encoded SWRL syntax and theoretical models can be integrated in software systems, specific for informed consent data management. In this work, we leverage the conceptual permission model from that previous study and SWRL encodings to complete information gaps for participants of a research study. We show a software implementation of our work, independent of an authoring environment such as Protégé, to show how reference ontologies in the legal health domain could be implemented in real-world software systems.

We used the theoretical proof of concept that was previously implemented in the Protege environment using four use case examples^9^. We provide a software-driven implementation to demonstrate the realization and feasibility of developing software to apply the permission model for health information technology. We postulate that permission model for informed consent could be realized for future health information technology and support health care interoperability. The output of the effort will provide a software implementation of the aforementioned sample use cases that can lead toward a minimally viable product for managing informed consent data and information.

## Methods

### Permission model

The permission model is a conceptualization of entities implicated when a human entity engages in a scientific research study. As a part of the informed consent life cycle, when an individual agrees to be a subject of a research study the act provides what is defined as a *consent directive* (http://purl.obolibrary.org/obo/ICO_0000322, an entity described in the Informed Consent Ontology) which “prescribes some action that is made allowable by a consenting entity.” With respect to HL7, it is a “legal record” for the participant which “permits or denies” entities. Essentially, when a participant expresses agreement to be in a study it “activates” authorization for connected entities. Some of the entities are *designated action* (http://purl.obolibrary.org/obo/ICO_0000324, what actions are allowable within the permission), *designated actor* (http://purl.obolibrary.org/obo/ICO_0000325, who is allowed as a result of the permission), and *designated object* (http://purl.obolibrary.org/obo/ICO_0000326, what are the materials permitted within the permission expression). **Figure *1*** summarizes the permission model with the association with the informed consent ontology.

**Figure 1.**
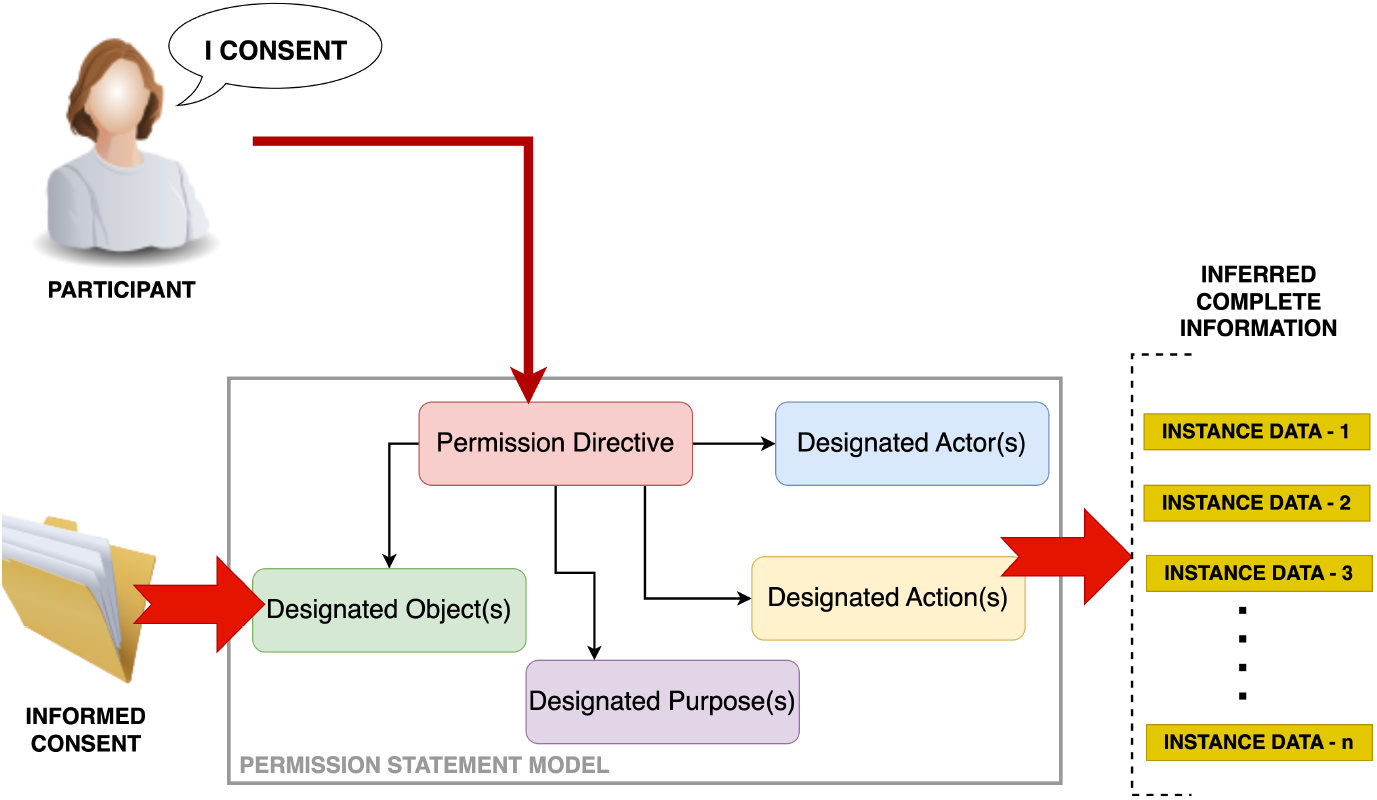
Description of the conceptual model for permissions using Informed Consent Ontology entities.

### Implementation of the permission model for informed consent

Researchers highlighted the theoretical implementation of the permission model for informed consent ^9^. Theoretically, an expression of consent is an instance manifested as a *OWLNamedIndividual*^11^. The real-world entities (actors, actions, objects, etc.) associated with the instance of the permission are represented as an instance. Using encoded Semantic Web Rule Language, we can generate additional instance data to complete entities not fully represented in the informed consent process that represents other entities that are not explicitly described.

### Summary of the software implementation

**Figure *2*** shows a summary of the software implementation of the execution of the simulation of capturing informed consent of the participant and the linking of entity data. A precondition of this is the encoding of the SWRL rules (see^9^ for the rules). Each rule was encoded using Protégé onto the informed consent ontology.

**Figure 2.**
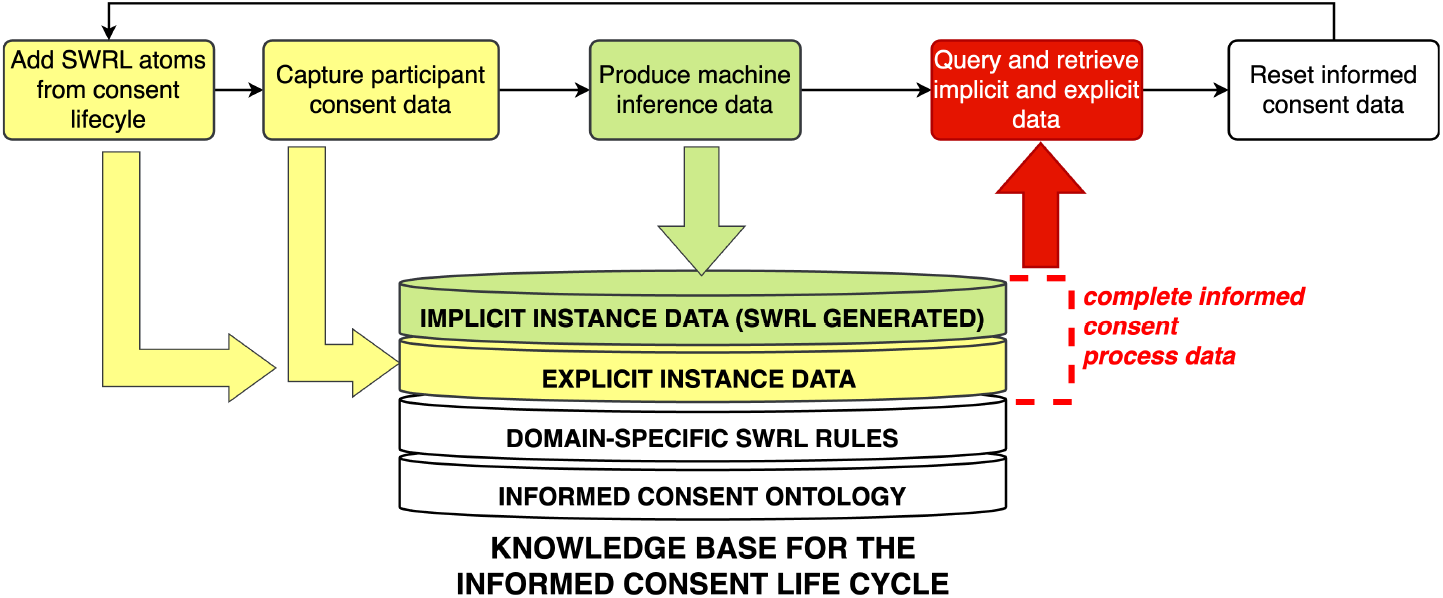
Process description of recording agreement and other informed consent data for application.

**Figure 3.**
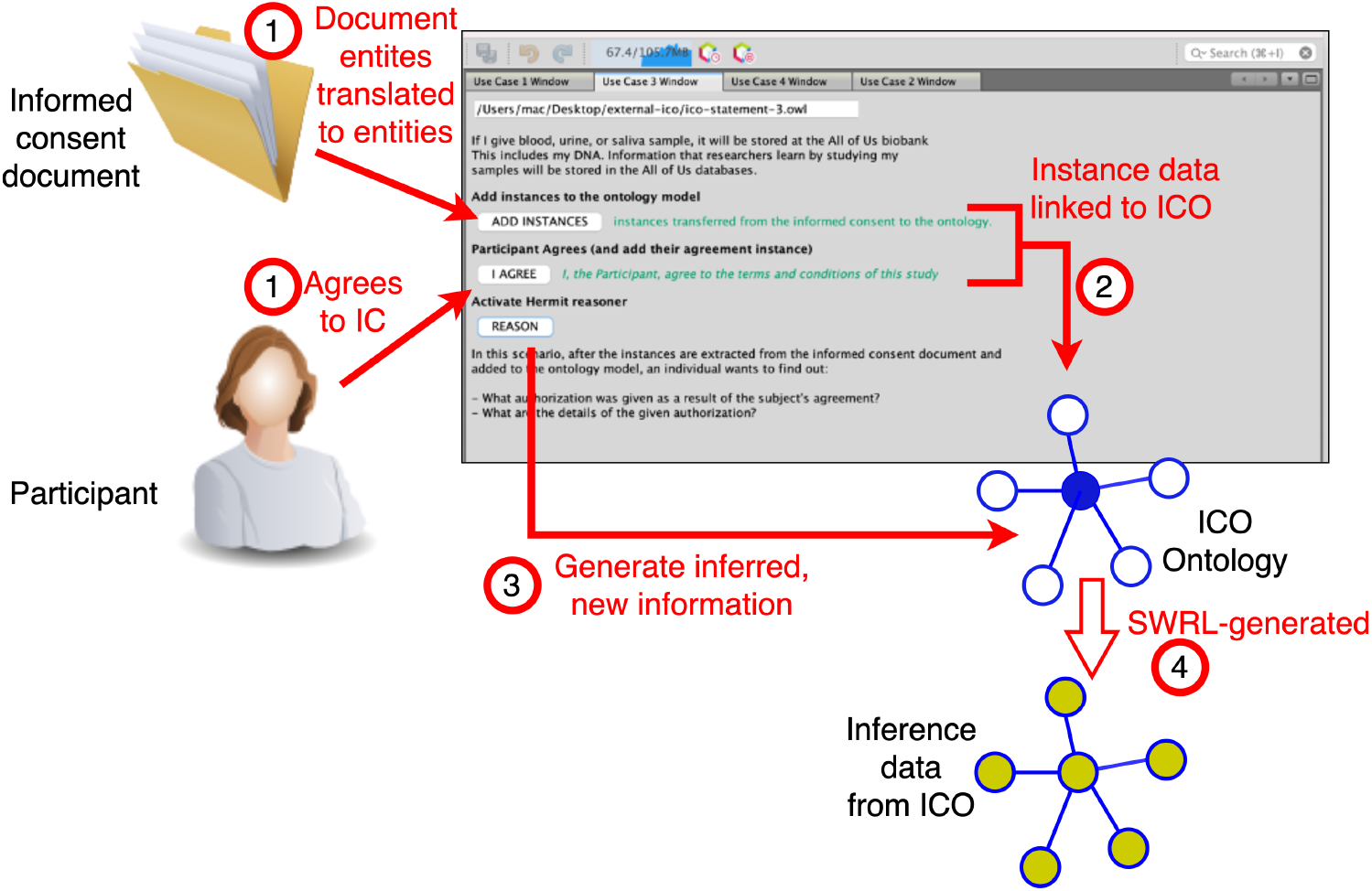
Operationalization of software implementation where entity data is linked to the Informed Consent Ontology and generates inferred data.

The general flow of the simulation presumes the individual participant digitally agrees (i.e., hypothetically captured through a health information technology tool for informed consent) to the participate. In addition, the SWRL rules provided in the previous study^9^ to generates missing/implicit information. On the software level, data about the informed consent process is recorded and captured, to be linked to the ontology model. The ICO model serves as an information schema. For example, “Martin Aerosmith” is the investigator of the study, an instance data of the type *designated principal investigator* (http://purl.obolibrary.org/obo/ICO_0000382). Similarly, if the study, for example, has biospecimen data as a part of the study, an instance data of type *designated biospecimen* (http://purl.obolibrary.org/obo/ICO_0000375) is linked to the ontology. The identification of all linked entities is to be identified by the informed consent documents or other resources.

With instance data corresponding from the informed consent lifecycle is linked to the ontology model, a software reasoner (e.g. HermiT^12^) performs the generation of implicit information based on SWRL rules and knowledge base of the ontology to provide complete informed consent information. The explicit and implicit data generated will be linked to the ontology model to allow for the querying. The HermiT reasoner provides an API to allow for programmatic reasoning from ontology models permitting for software integration. While discussion of ontology model reasoning is beyond the scope of this piece, essentially, a reasoner (e.g., FaCT++, HermiT, Pellet, Elk, etc.) performs tests of *classification* based on the axioms (which includes the SWRL rules) and *coherency* of the expression of axioms. By extension, it is able to describe implicit information from the axioms. With the implicit information generated and linked to the ontology, we can easily use SPARQL or other programmatic means to retrieve information from the ontology model.

## Results

We developed a Java software application that simulates the implementation of the conceptual permission model using the Informed Consent Ontology. The goal of this tool is to demonstrate an information technology translation of the methodology for real-world use. We used the NetBeans Rich Client Platform^13^, a Java-based software development application framework to create software simulation for the capturing and processing informed consent data from a participant. The application simulates sample four use cases of the All of Us informed consent, and integrates the Informed Consent Ontology (ICO).

### First permission use case

*Please check the box below if you agree to take part: I have read this consent form (or someone read it to me). I understand the information in this form. All of my questions have been answered. I freely and willingly choose to take part in the All of Us Research Program*.

The first use case involves the participant agreeing and understanding that he/she was informed by the research team. The participant had submitted an agreement and giving designated researchers permission. This also implies that detailed explanation was provided at the time of consent.

We show the results of the implementation (**Figure *4*** and **Figure *5***) to simulate the execution of the conceptual permissions and the production of inferred information. With **Figure *4***, the instance data produces inference links between the primary investigator and the investigative team (*_pi* and *_team*, respectively) with the instance data associated with explaining the details of the research protocol (*_explained*). The primary investigator and the research team were automatically classified as *designated permitted actor* (http://purl.obolibrary.org/obo/ICO_0000378) to indicate these entities of the life cycle are permitted to proceed in the study. The instances of explanation of the protocol was auto-categorized as *explaining benefits of participation to participant candidate* (http://purl.obolibrary.org/obo/ICO_0000121), *explaining record confidentiality to participant candidate* (http://purl.obolibrary.org/obo/ICO_0000135), *explaining risks of participation to participant candidate* (http://purl.obolibrary.org/obo/ICO_0000128), *explaining study protocol to participant candidate* (http://purl.obolibrary.org/obo/ICO_0000117), and *explaining study purpose to participant candidate* (http://purl.obolibrary.org/obo/ICO_0000105) to indicate prescribed details of what (and should be) was communicated to the participants. In **Figure *5***, which is part of the first use case, data was recorded of the subject’s approval to be involved in the hypothetical study. This is expressed with an inference link (Relation Ontology’s *participates in*, i.e., http://purl.obolibrary.org/obo/RO_0000056 and *has participants*, i.e., http://purl.obolibrary.org/obo/RO_0000057) connecting the entity instance data of the subject with the instance data of type *legally effective consenting* (http://purl.obolibrary.org/obo/ICO_0000142) for representing agreement to the informed consent. Overall, this allowed us to inquire what exactly was explained to the participant (“*What was explained to the subject?*’’) and who explained the details of the protocol (2 of **Figure *4***) and whether the participant gave them consent to proceed with the study (3 of **Figure *5***).

**Figure 4.**
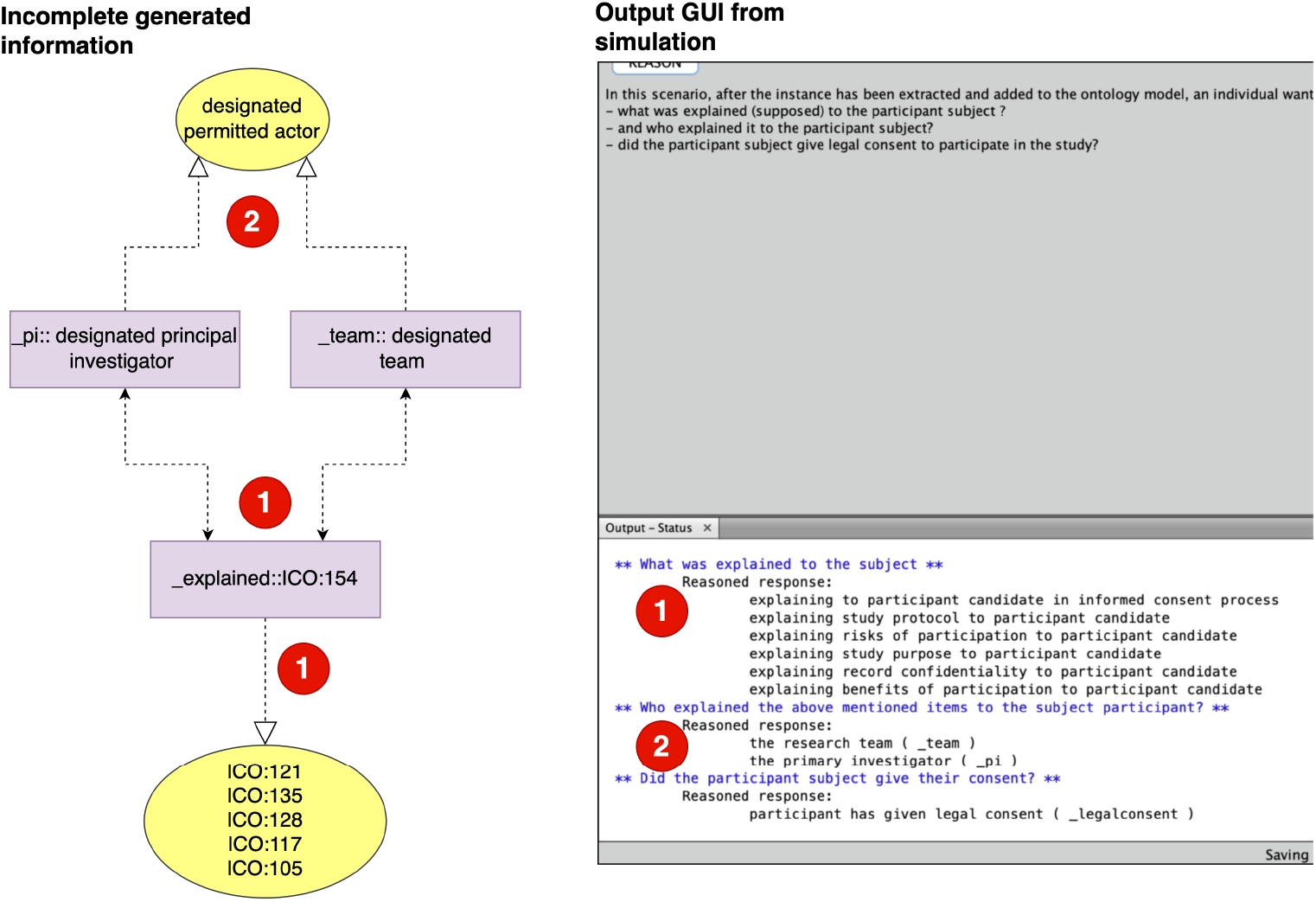
First use case implemented within the software application. Dotted lines de notes labeled inferenced links (object properties) and yellow ovals denotes inferred classification, by way of the SWRL encodings in the Informed Consent Ontology.

**Figure 5.**
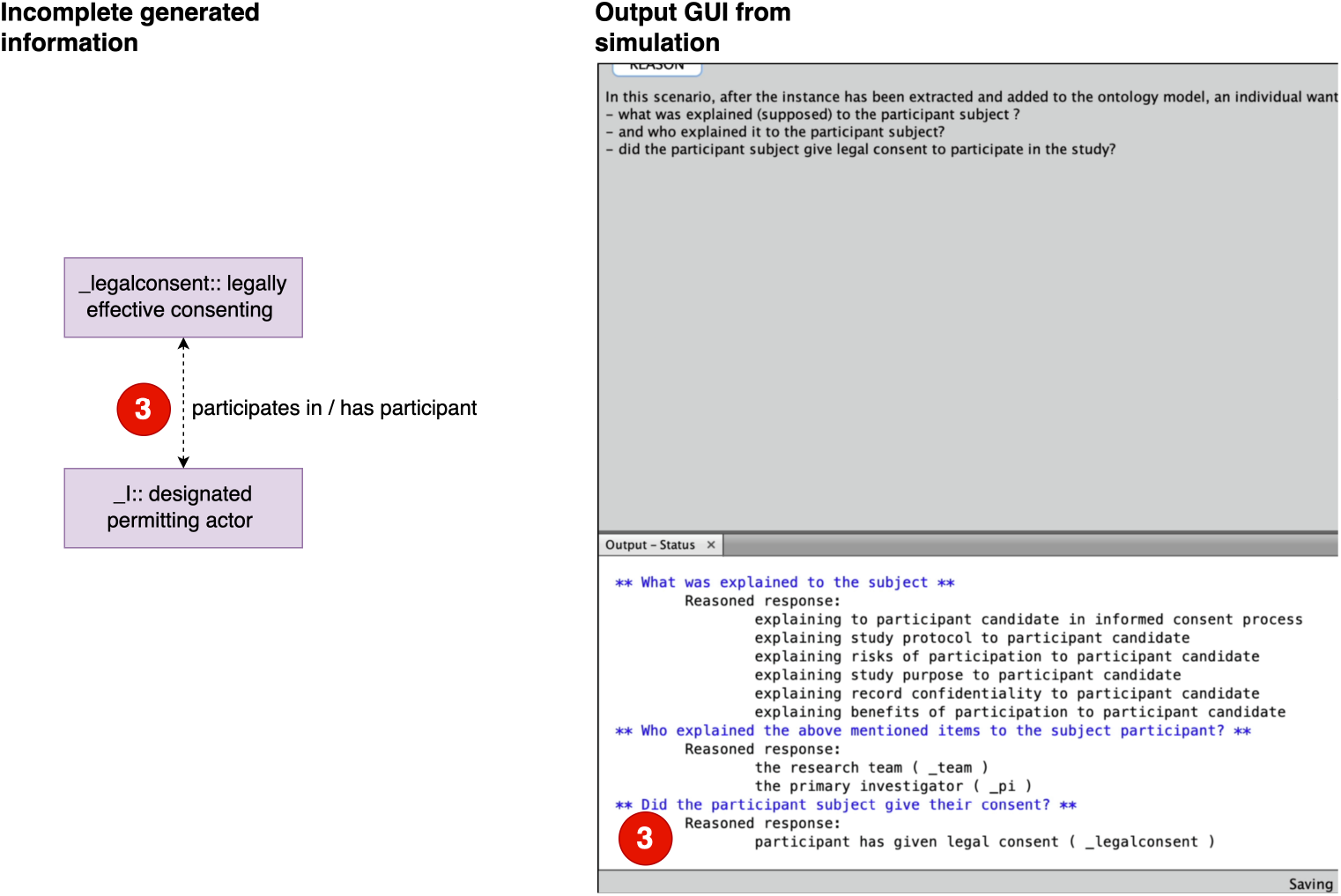
First use case implemented within the software application. Dotted lines denotes labeled inferenced links (object properties)

### Second permission use case

*Results that might change your medical care. These are results that could be used by a healthcare provider to take better care of you. For example, if any of your physical measurements are outside of what we would expect, we will tell you so you can follow-up with your healthcare provider. You will have to pay for the cost of follow-up care with your own healthcare provider*

The second use case recommends the participant to seek medical care if any potential issues were to result of being involved in the study. This rule implies that said obligation of the participant to find medical treatment, and that this information was delivered by the research team. **Figure *6*** demonstrate the implementation of the second use case, similar to the previous case. The reasoner was able to execute the SWRL rules that expressed an inferenced link (Relation Ontology’s *participates in* and *has participants*) between the instance of the participant and the instance of *standard medical treatment* (http://purl.obolibrary.org/obo/ICO_0000110) to indicate the responsibility of the participant to seek medical treatment. Also, the responsibility that the team and the primary investigator should have communicated this information. This was expressed with an inferenced link connecting the team and primary investigator data instance entity with the instance of the *act of communicating* (http://purl.obolibrary.org/obo/ICO_0000269). The produced inferenced allowed for an inquiry about what is the responsibility of the participant (1 of **Figure *6***) and who informed the participant of this responsibility (2 of **Figure *6***).

**Figure 6.**
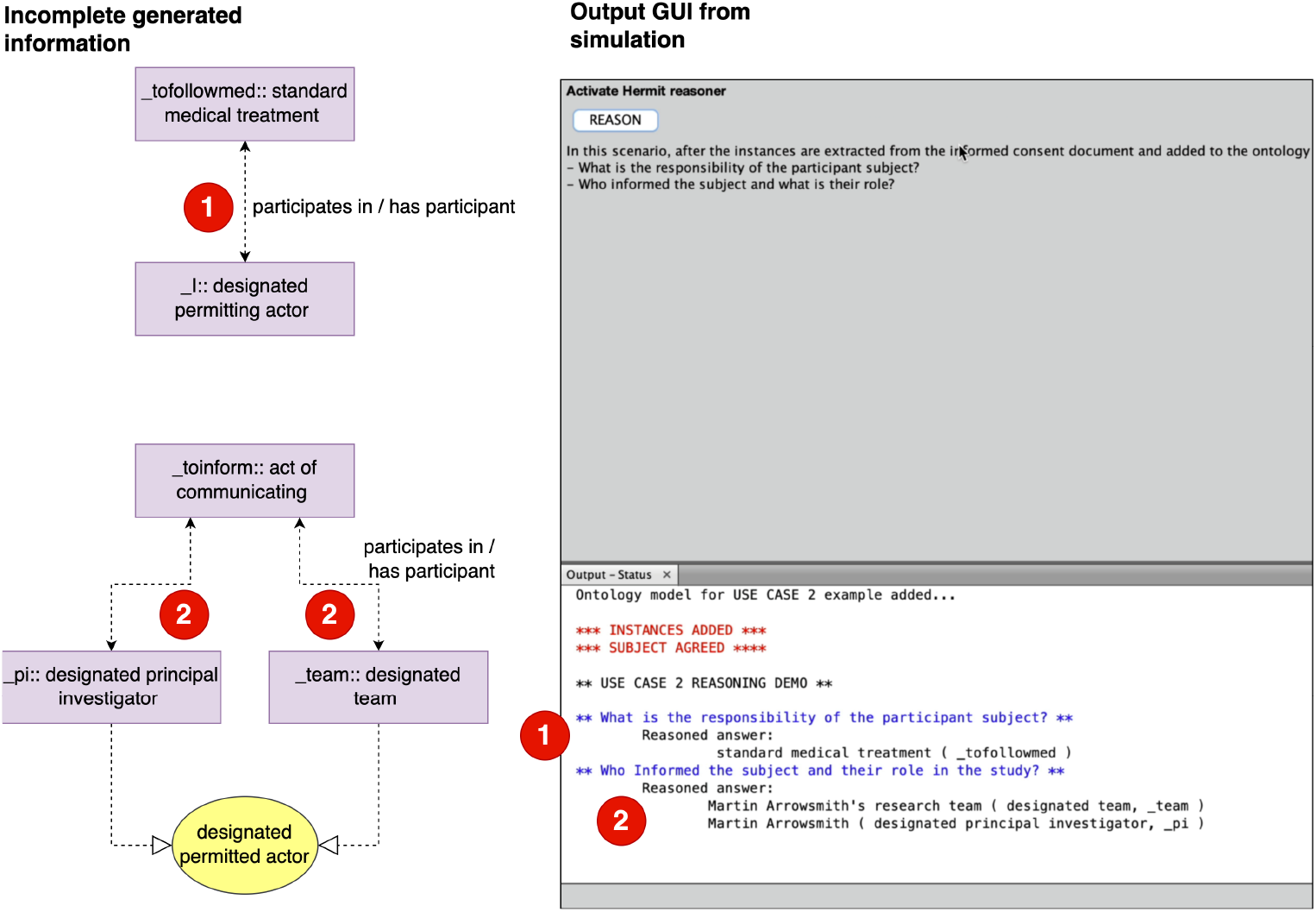
Second use case implemented within the software application. Dotted lines denotes labeled inferenced links (object properties) and yellow ovals denotes inferred classification, by way of the SWRL encodings in the Informed Consent Ontology.

### Third and fourth permission use case

*If I give blood, urine, or saliva sample, it will be stored at the All of Us biobank. This includes my DNA. Information that researchers learn by studying my samples will be stored in the All of Us databases*

*Researchers will do studies using the All of Us databases and biobank. Their research may be on nearly any topic*

The third and fourth use case involves permissions to store and share bio-specimen data from the participant and to subsequently use the data for secondary research analysis. The third use case implies that collected bio-specimen data has allowed for data sharing and storing, and have given their authorization for storing and sharing bio-specimen data. It also implicitly denotes that the act of storing and sharing is only for *urine, blood*, and *saliva*. The fourth use case implies the researchers who are designated to the study have only this authorization.

**Figure *7*** and **Figure *8*** displays the execution of the third and fourth use case. As a result of agreeing, an authorization instance (*_authorize* of type of *act of authorizing* (http://purl.obolibrary.org/obo/ICO_0000046)) links, by way of inference from SWRL rule to instances of the *act of storing a specimen* (http://purl.obolibrary.org/obo/ICO_0000060) (\*_tosharedata* of **Figure *7***), the *act of data sharing* (http://purl.obolibrary.org/obo/ICO_0000228) (*_tosharedata*), and the *act of using study participating data* (http://purl.obolibrary.org/obo/ICO_0000116) (*_tosusedata* of **Figure *8***). The instances of *designated bio-specimen* (http://purl.obolibrary.org/obo/ICO_0000375) (*blood, urine*, and *saliva*) have inferred links (Data Use Ontology’s *is restricted to*, i.e., http://purl.obolibrary.org/obo/DUO_0000010) to the aforementioned instances of *act of storing a specimen* (http://purl.obolibrary.org/obo/ICO_0000060) and *act of using study participant data* (http://purl.obolibrary.org/obo/ICO_0000116). The SWRL rule generated object properties (“links”) for Relation Ontology’s *participates in* and *has participants*. Both the primary investigator and the investigative team (*_pi* and *_team*, 3 of **Figure *7*** and 3 of **Figure *8***) have inferred links to storing and act of using study participant data to indicate permitted individuals who can participate in these actions.

**Figure 7.**
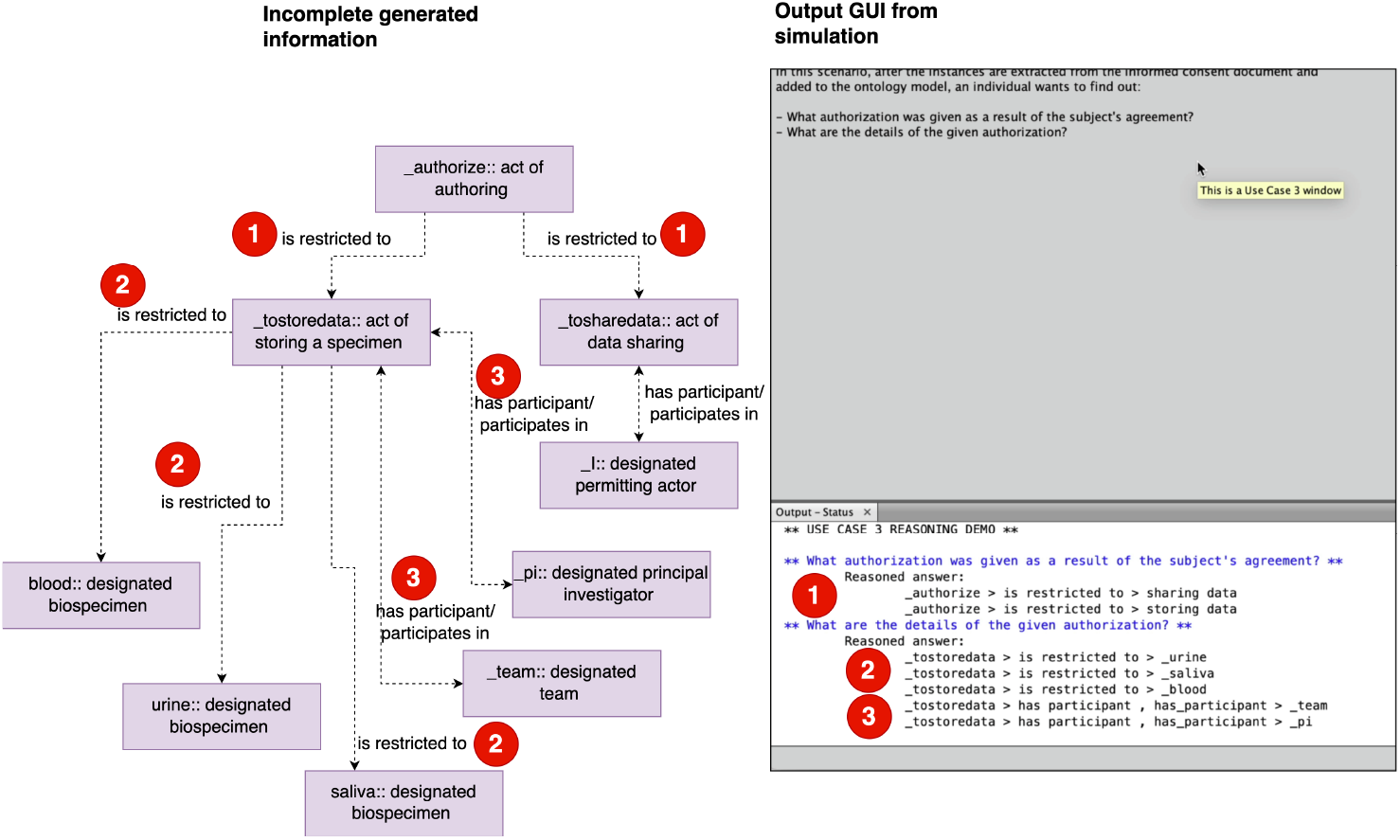
Third use case implemented within the software application. Dotted lines de- notes labeled inferenced links (object properties)

**Figure 8.**
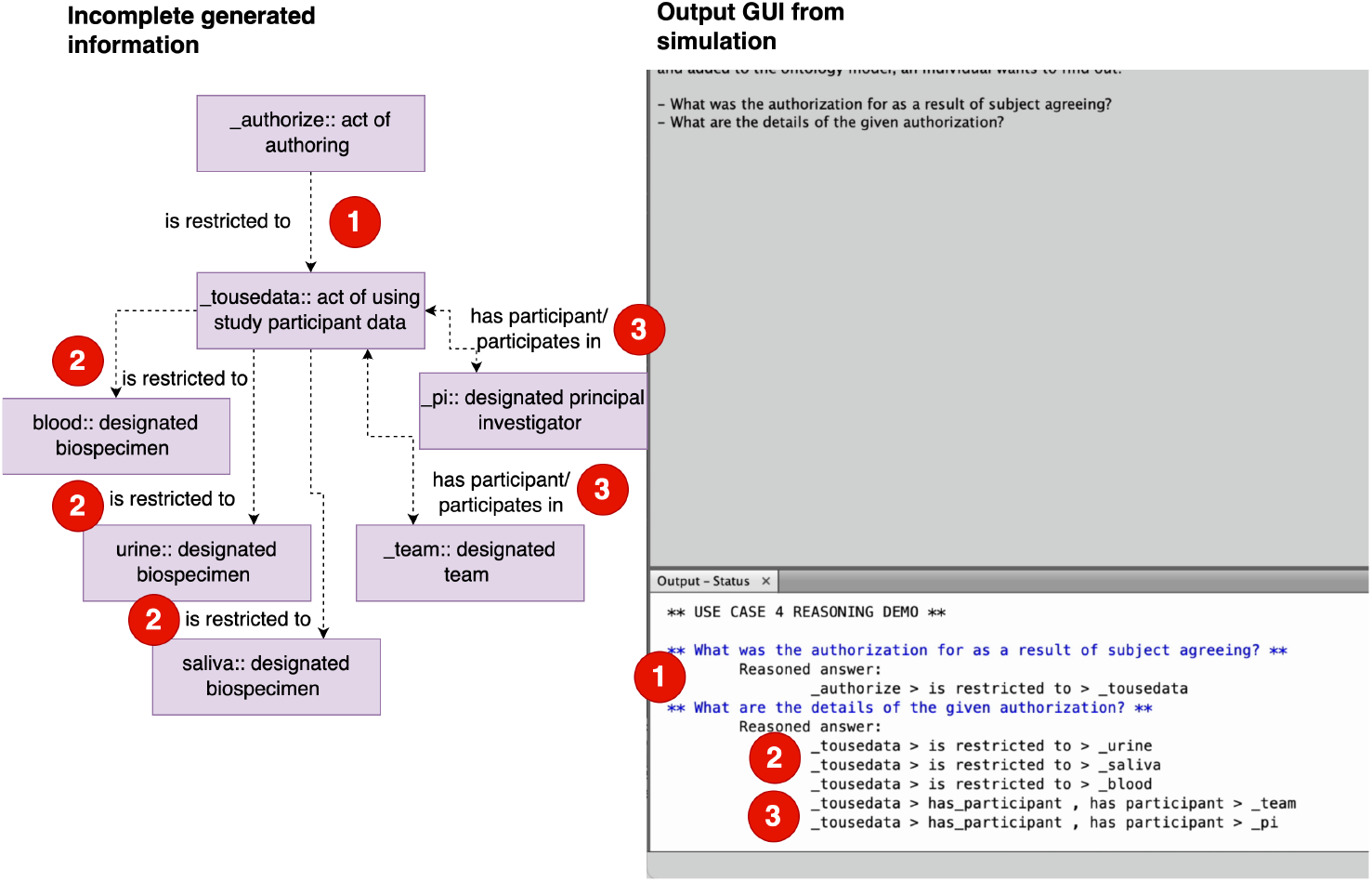
Fourth use case implemented within the software application. Dotted lines denotes labeled inferenced links (object properties).

Our work which includes the sample ontology models that have the encoded domain SWRL rules is available on our GitHub repository at https://github.com/ProfTuan/ICO-Integration-Application-Demo. The software developed with Java 11 using NetBeans, and the sample models demonstrated in this study are encoded in OWL2. All materials are released under the Creative Commons license.

## Discussion and Conclusions

We presented an applied exhibition of how we implemented conceptual permission models for informed consent entities for future software system application, specifically for health care. Leveraging the informed consent ontology that is coupled with Semantic Web Rule Language (SWRL) syntax that encodes and represents permissions of the participant consent, we showed how the ontology can integrate and execute its intrinsic knowledge base with software code to simulate the implementation of linking and describing and expressing implicit (*i*.*e*., incomplete) information of entities within the life cycle of informed consent process. This work also extends previous research that described a proof of concept method for translational application in health care and research. Overall, this further shows the premise of how a reference ontology for informed consent could be applied in real world use case.

Collecting informed consent electronically is a well investigated area^14–16^. However, none of the documented research on this area considers or utilize resources to facilitate data integration, such as ontologies or controlled terminologies to assist. Health information silos is an ongoing issue in health information systems, and there is an opportunity to employ terminological standards (i.e., ontologies) to support this effort. In addition, this effort realizes a unique method in using computable ontologies to provide comprehensive and tailored informed consent information to consumers (participants, clinicians, etc.).

Our future work will be directed on informed consent for vaccine research. In the United States as well as other regions around the world, informed consent is required prior to vaccine inoculation by a health care provider. Previous researchers have extended the Informed Consent Ontology for vaccines (e.g. Vaccine Informed Consent Ontology). The Vaccine Informed Consent Ontology (VICO) represents extended entities for vaccination^17^. Some examples include entities like *intranasal influenza vaccination* (VICO_0000043) and *vaccination informed consent form* (ICO_0000138), where former is a type of *vaccination planned process* “that occurs in intranasal route” and the latter is “an informed consent form that is used for vaccination”.

With foundational work, we intend to explore the possibility of applying the methods and implementation from this study for vaccine informed consent domain. The informed consent in vaccination domain shares some of the legal, ethical issues present within the informed consent life cycle. Presumably if we were to extend the previous work in vaccine informed consent, we could mitigate the knowledge and information gap that sometimes impedes vaccination rates, as well as showing relevance in using the Informed Consent Ontology (ICO) in integrative software applications for health care and scientific research.

Following methodology from previous researchers, we developed an executable implementation of conceptual permissions derived from a reference ontology for informed consent (Informed Consent Ontology). The outcome of this work demonstrated potential applied implementation of a conceptual framework for permissions to capture participant agreement and manage data from informed consent lifecycle. This agreement data enacted links to simulated data associated with informed consent entities, including producing inferenced missing information. The translational outcome could encourage the use of the Informed Consent Ontology for software applications that need to harmonize data cross different platforms, thereby, furthering the goals of interoperability of health care systems.

Future direction will explore unique sub-domain of vaccine informed consent toward the development of tools and technology supported by reference ontologies like Informed Consent Ontology.

## Data Availability

All data produced are available online at GitHub

https://github.com/ProfTuan/ICO-Integration-Application-Demo

## Acknowledgements

This research was supported by the NIH/NCI under Contract Number 75N91020C00017 (Manion PI), and builds off work supported by an award from the Michigan Institute for Data Science (Harris PI), and NIH/NHGRI U01 HG009454 (Tao PI); by the National Institute of Allergy and Infectious Diseases of the National Institutes of Health under Award No, R01AI130460 (Tao, PI), R01AI130460-03S1 (Tao, PI), and U24AI171008 (He and Tao, MPIs); by the National Human Genome Research Institute under Award No, R03HG013341; and by the Cancer Prevention Research Institute of Texas under Award No, #RP220244.

